# Forecasting COVID-19 impact on hospital bed-days, ICU-days, ventilator-days and deaths by US state in the next 4 months

**DOI:** 10.1101/2020.03.27.20043752

**Authors:** IHME COVID-19 health service utilization forecasting team, Christopher JL Murray

## Abstract

**Importance:** This study presents the first set of estimates of predicted health service utilization and deaths due to COVID-19 by day for the next 4 months for each state in the US.

**Objective:** To determine the extent and timing of deaths and excess demand for hospital services due to COVID-19 in the US.

**Design, Setting, and Participants:** This study used data on confirmed COVID-19 deaths by day from WHO websites and local and national governments; data on hospital capacity and utilization for US states; and observed COVID-19 utilization data from select locations to develop a statistical model forecasting deaths and hospital utilization against capacity by state for the US over the next 4 months.

**Exposure(s):** COVID-19.

**Main outcome(s) and measure(s):** Deaths, bed and ICU occupancy, and ventilator use.

**Results:** Compared to licensed capacity and average annual occupancy rates, excess demand from COVID-19 at the peak of the pandemic in the second week of April is predicted to be 64,175 (95% UI 7,977 to 251,059) total beds and 17,380 (95% UI 2,432 to 57,955) ICU beds. At the peak of the pandemic, ventilator use is predicted to be 19,481 (95% UI 9,767 to 39,674). The date of peak excess demand by state varies from the second week of April through May. We estimate that there will be a total of 81,114 (95% UI 38,242 to 162,106) deaths from COVID-19 over the next 4 months in the US. Deaths from COVID-19 are estimated to drop below 10 deaths per day between May 31 and June 6.

**Conclusions and Relevance:** In addition to a large number of deaths from COVID-19, the epidemic in the US will place a load well beyond the current capacity of hospitals to manage, especially for ICU care. These estimates can help inform the development and implementation of strategies to mitigate this gap, including reducing non-COVID-19 demand for services and temporarily increasing system capacity. These are urgently needed given that peak volumes are estimated to be only three weeks away. The estimated excess demand on hospital systems is predicated on the enactment of social distancing measures in all states that have not done so already within the next week and maintenance of these measures throughout the epidemic, emphasizing the importance of implementing, enforcing, and maintaining these measures to mitigate hospital system overload and prevent deaths.

**Data availability statement:** A full list of data citations are available by contacting the corresponding author.

**Funding Statement:** Bill & Melinda Gates Foundation and the State of Washington

**Key Points:** *Question:* Assuming social distancing measures are maintained, what are the forecasted gaps in available health service resources and number of deaths from the COVID-19 pandemic for each state in the United States?

*Findings:* Using a statistical model, we predict excess demand will be 64,175 (95% UI 7,977 to 251,059) total beds and 17,380 (95% UI 2,432 to 57,955) ICU beds at the peak of COVID-19. Peak ventilator use is predicted to be 19,481 (95% UI 9,767 to 39,674) ventilators. Peak demand will be in the second week of April. We estimate 81,114 (95% UI 38,242 to 162,106) deaths in the United States from COVID-19 over the next 4 months.

*Meaning:* Even with social distancing measures enacted and sustained, the peak demand for hospital services due to the COVID-19 pandemic is likely going to exceed capacity substantially. Alongside the implementation and enforcement of social distancing measures, there is an urgent need to develop and implement plans to reduce non-COVID-19 demand for and temporarily increase capacity of health facilities.

## Background

The Coronavirus Disease 2019 (COVID-19) pandemic started in Wuhan, China, in December 2019^1^ and has since spread to the vast majority of countries.^2^ As of March 24, 5 countries have recorded more than a thousand deaths: China, France, Iran, Italy, and Spain. COVID-19 is not only causing mortality but is also putting considerable stress on health systems with large case numbers. In the US, COVID-19 has spread to all 50 states, with 31 states reporting deaths so far. Estimates of the potential magnitude of COVID-19 patient volume are urgently needed for US hospitals to effectively manage the rising case load and provide the highest quality of care possible.

COVID-19 forecasts have largely been based on mathematical models that capture the probability of moving between states from susceptible to infected, and then to a recovered state or death (SIR models). Many SIR models have been published or posted online.^3–20^ In general, these models assume random mixing between all individuals in a given population. While results of these models are sensitive to starting assumptions and thus differ between models considerably, they generally suggest that given current estimates of the basic reproductive rate (the number of cases caused by each case in a susceptible population), 25% to 70% of the population will eventually become infected.^6,20^ Based on reported case-fatality rates, these projections imply that there would be millions of deaths in the United States due to COVID-19. However, individual behavioral responses and government-mandated social distancing (school closures, non-essential service closures, and shelter-in-place orders) can dramatically influence the course of the epidemic. In Wuhan, strict social distancing was instituted on January 23, 2020, and by the time new infections reached 1 or fewer a day (March 15, 2020), the confirmed proportion of the population infected was less than 0.5%. SIR models with assumptions of random mixing can overestimate health service need by not taking into account behavioral change and government-mandated action. Using reported case numbers and models based on those for health service planning is also not ideal because of widely varying COVID-19 testing rates and strategies. For example, South Korea has undertaken aggressive population-based screening and testing, while in the US, limited test availability has led to largely restricting testing to those with more severe disease or those who are at risk of serious complications.

An alternative strategy is to focus on modeling the empirically observed COVID-19 population death rate curves, which directly reflect both the transmission of the virus and the case-fatality rates in each community. Deaths are likely more accurately reported than cases in settings with limited testing capacity where tests are usually prioritized for the more severely ill patients. Hospital service need is likely going to be highly correlated with deaths, given predictable disease progression probabilities by age for severe cases. In this study, we use statistical modeling to implement this approach and derive state-specific forecasts with uncertainty for deaths and for health service resource needs and compare these to available resources in the US.

## Methods

The modeling approach in this study is divided into four components: (i) identification and processing of COVID-19 data; (ii) statistical model estimation for population death rates as a function of time since the death rate exceeds a threshold in a location; (iii) predicting time to exceed a given population death threshold in states early in the pandemic; and (iv) modeling health service utilization as a function of deaths.

### Data identification and processing

Local government, national government, and WHO websites^21–25^ were used to identify data on confirmed COVID-19 deaths by day at the first administrative level (state or province, hereafter “admin 1”). Government declarations were used to identify the day different jurisdictions implemented various social distancing policies (stay-at-home or shelter-in-place orders, school closures, closures of non-essential services focused on bars and restaurants, and the deployment of severe travel restrictions) following the New Zealand government schema.^26^ Data on timings of interventions were compiled by checking national and state governmental websites, executive orders, and newly initiated COVID-19 laws. Data on licensed bed and ICU capacity and average annual utilization by state were obtained from the American Hospital Association.^27^ We estimated ICU utilization rates by multiplying total bed utilization rates by the ratio of ICU bed utilization rates over total bed utilization rates from a published study.^28^ Observed COVID-19 utilization data were obtained for Italy^21^ and the United States,^29^ providing information on inpatient and ICU use. Data from China^30^ were used to approximate inpatient and ICU use by assuming that severe patients were hospitalized and critical patients required an ICU stay. Other parameters were sourced from the scientific literature and an analysis of available patient data.^31^ Age-specific data on the relative population death rate by age are available from China,^30^ Italy,^32^ Korea,^33^ and the US^29^ and show a strong relationship with age (Figure 1).

**Figure 1.**
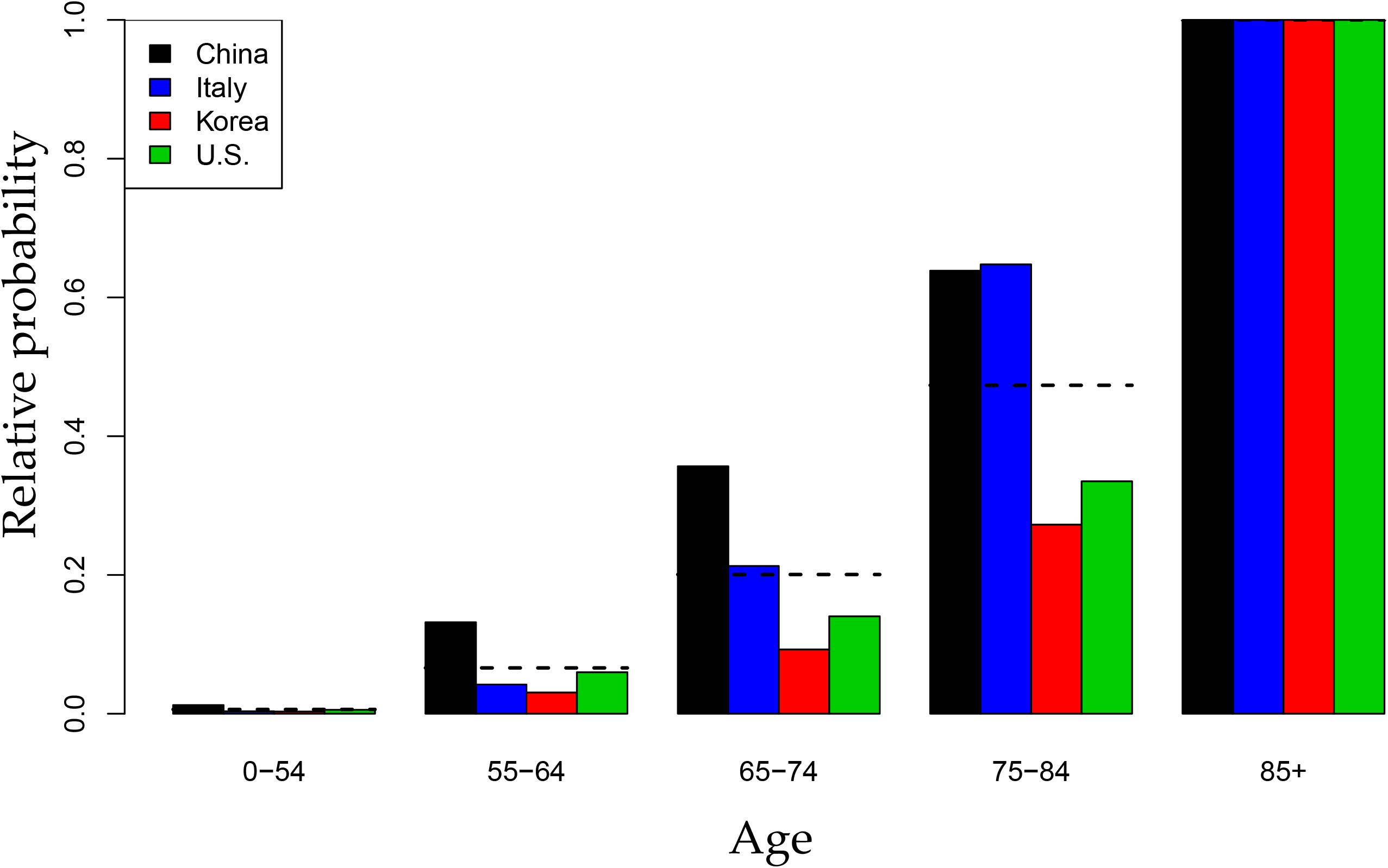
Normalized age-pattern of death based on data from Italy, South Korea, China, and the US

Using the average observed relationship between the population death rate and age, data from different locations can be standardized to the age structure using indirect standardization. For the estimation of statistical models for the population death rate, only admin 1 locations with an observed death rate greater than 0.31 per million (e-15) were used. This threshold was selected by testing which threshold minimized the variance of the slope of the death rate across locations in subsequent days.

A covariate of days with expected exponential growth in the cumulative death rate was created using information on the number of days after the death rate exceeded 0.31 per million to the day when 4 different social distancing measures were mandated by local and national government: school closures, non-essential business closures including bars and restaurants, stay-at-home recommendations, and travel restrictions including public transport closures. Days with 1 measure were counted as 0.67 equivalents, days with 2 measures as 0.334 equivalents and with 3 or 4 measures as 0. For states that have not yet implemented all of the closure measures, we assumed that the remaining measures will be put in place within 1 week. This lag between reaching a threshold death rate and implementing more aggressive social distancing was combined with the observed period of exponential growth in the cumulative death rate seen in Wuhan after Level 4 social distancing was implemented, adjusted for the median time from incidence to death. For ease of interpretation of statistical coefficients, this covariate was normalized so the value for Wuhan was 1.

### Statistical model for the cumulative death rate

We developed a curve-fitting tool to fit a nonlinear mixed effects model to the available admin 1 cumulative death data. The cumulative death rate for each location is assumed to follow a parametrized Gaussian error function:

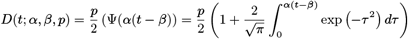

where the function ψ is the Gaussian error function (written explicitly above), *p* controls the maximum death rate at each location, *t* is the time since death rate exceeded 1e-15, *β* (beta) is a location-specific inflection point (time at which rate of increase of the death rate is maximum), and α (alpha) is a location-specific growth parameter. Other sigmoidal functional forms (alternatives to ψ) were considered but did not fit the data as well. Data were fit to the log of the death rate in the available data, using an optimization framework described in the appendix.

We ensembled two types of models to produce the estimates. We either parametrized the level parameter p or the time-axis shift parameter beta to depend on a covariate based on time from when the initial death rate exceeds 1e-15 to the implementation of social distancing. The value of the covariate multipliers in each type of model was assumed to closely follow the fit obtained from data from Wuhan, which is the time series to reach a stable state in the training dataset. To be specific, the generalizable information from Wuhan was the impact that social distancing had on maximum death rate and time to reach the inflection point. For each type of model, we both considered ‘short-range’ and ‘long-range’ variants, to explain existing data and forecast long- term trends, respectively. In the former case, covariate multipliers could deviate from those fit to Wuhan, while in the latter, the data from Wuhan had a larger impact on the final covariate multiplier. At the draw level, we linearly interpolate between the models as we go from times where we have collected data already to long-term forecasts. Specifically, we take the weighted combination of the daily increment of the log death rates from these models, with the weight linearly transitioning from short-range to long-range. The two remaining parameters (not modeled using covariates) were allowed to vary among locations to explain location-specific data.

Uncertainty in the model estimates is driven by two components: (1) uncertainty from fixed effect estimation and (2) uncertainty from random effects, with the latter dominant because of the high variation between locations. Uncertainty of fixed effects is estimated using asymptotic statistics derived from the likelihood. In every model, we estimated location-specific parameters or multipliers, and therefore used the empirical variance-covariance matrix of these parameters as a prior for location-specific fit. Posterior uncertainty within each location was then obtained using a standard asymptotic approximation at that location. Once we estimated total uncertainty by location at the draw level, overall uncertainty was then obtained by aggregation of draws. The dataset age-standardized to the age-structure of California is shown in Figure 2.

**Figure 2.**
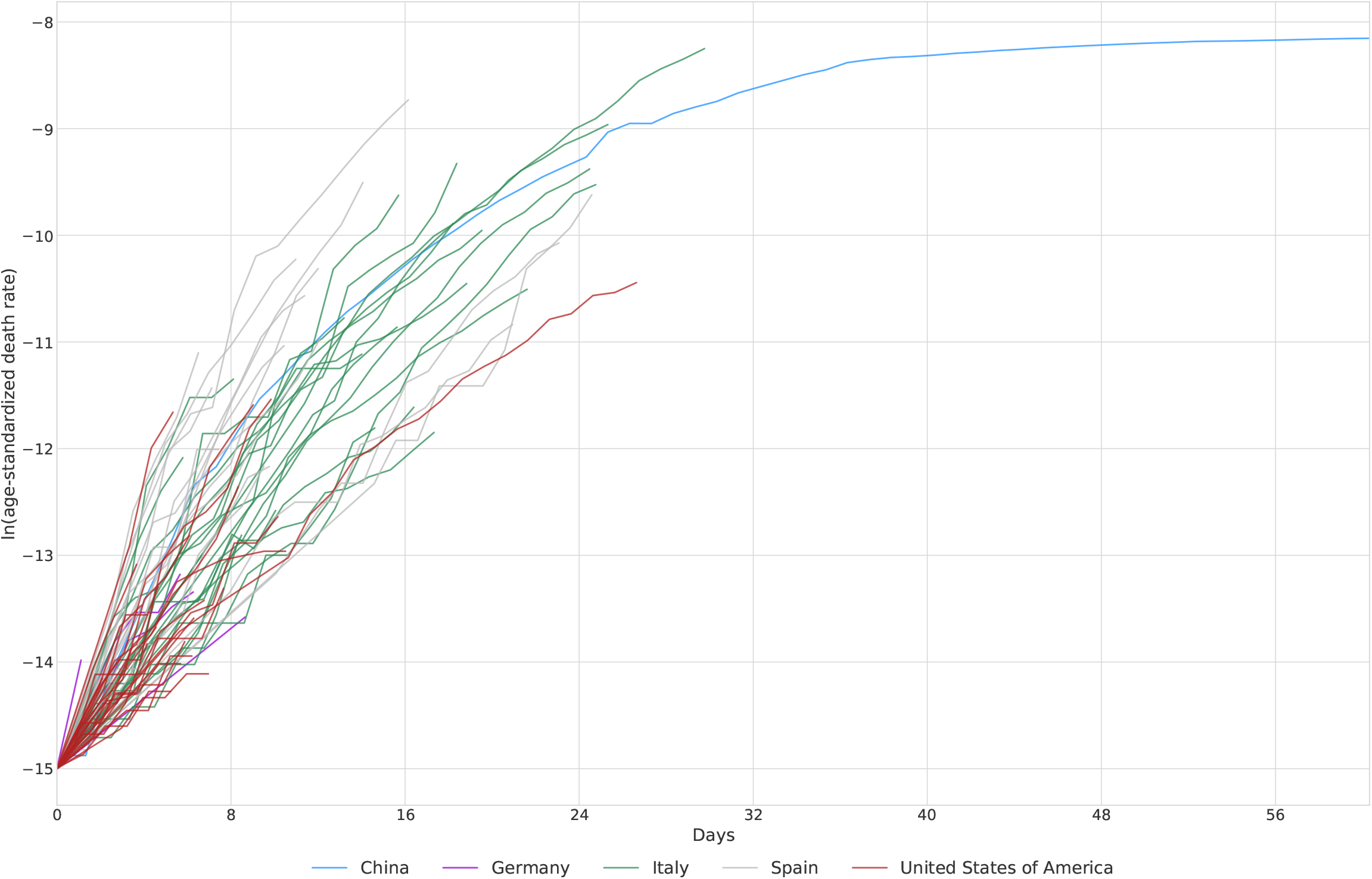
Death rate data age-standardized to California as a function of time since a threshold death rate of 0.3 per million.

Because of the unique high-intensity epidemic in the Life Care Kirkland facility in Washington state,^34,35^ we have modeled this facility separately from the general population – see the appendix for details. Furthermore, as our initial development of the model was focused on King and Snohomish counties in Washington state, we have also stratified these 2 counties from the rest of Washington state. In other words, for Washington state, we model 3 populations explicitly: (i) the Life Care Kirkland facility; (ii) the remainder of the King and Snohomish county population; and (iii) all other counties in Washington state.

### Time to threshold death rate

Only 27 states have deaths greater than 0.31 per million (e-15) and were included in the model estimation along with data on 44 other admin 1 locations. For other US states, we estimated the expected time from the current case count to reach the threshold level for the population death rate model. Using the observed distribution of the time from each level of case count to the threshold death rate for all admin 1 locations with data, we estimated this distribution. We used the mean and standard deviation of days from a given case count to the threshold death rate to develop the probability distribution for the day each state will cross over the threshold death rate, and then we applied the death rate epidemic curve after crossing the threshold.

### Hospital service utilization microsimulation model

From the projected death rates, we estimated hospital service utilization using an individual-level microsimulation model. We simulated deaths by age using the average age pattern from Italy, China, South Korea, and the US (Figure 1) due to the relatively small number of deaths included for the US (n = 46) and the fact that the US age pattern is likely biased toward older-age deaths due to the early nursing home outbreak in Washington state. For each simulated death, we estimated the date of admission using the median length of stay for deaths estimated from the global line list (10 days <75 years; 8 days 75+ years). Simulated individuals requiring admission who were discharged alive were generated using the age-specific ratio of admissions to death (Figure 3), based again on the average across Italy, China, and the US. The age-specific fraction of admissions requiring ICU care was based on data from the US (122 total ICU admissions over 509 total admissions). The fraction of ICU admissions requiring invasive ventilation was estimated as 54% (total n = 104) based on 2 studies from China.^36,37^ To determine daily bed and ICU occupancy and ventilator use, we applied median lengths of stay of 12 days based on the analysis of available unit record data and 8 days for those admissions with ICU care.^37^

**Figure 3.**
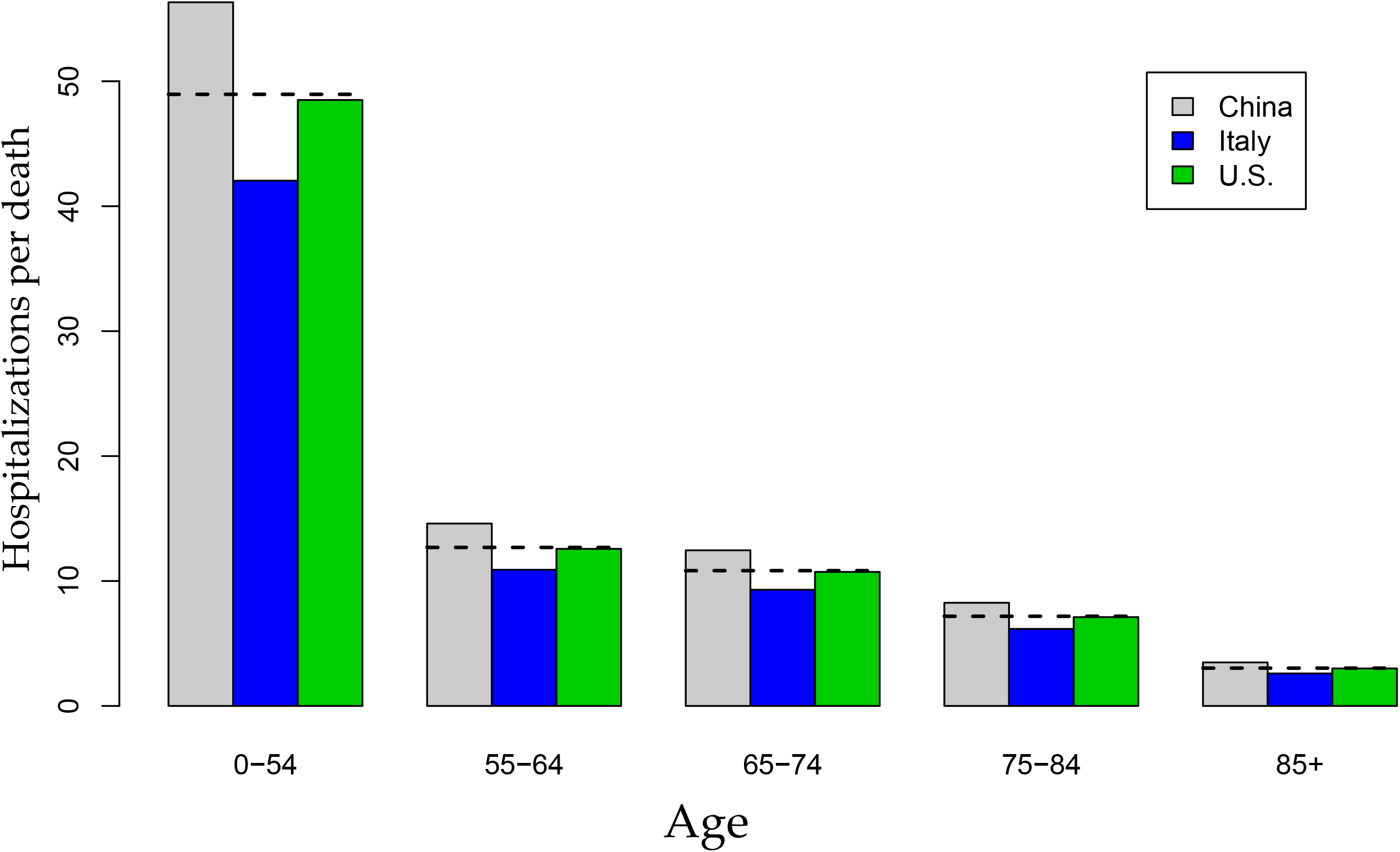
Age-specific ratio of admissions to deaths based on data from Italy, China, and the US

## Results

By aggregating forecasts across states, we determined the overall trajectory of expected health care need in different categories and deaths, as shown in Figure 4. Demand for health services rapidly increases in the last week of March and first 2 weeks of April and then slowly declines through the rest of April and May, with demand continuing well into June. The shape of the curve reflects both the epidemic curves within each state and the staggered nature of the epidemic around the country. Daily deaths in the mean forecast exceed 2,300 by the second week of April. While peak demand will occur at the national level in the second week of April, this varies by state as shown in Figure 5. Peak demand will occur in the first half of April in about a third of states. This includes states like New York which have had early epidemics and a corresponding sharp rise in deaths. In contrast, other states such as Washington State and California with early epidemics have experienced slower increases in deaths.

**Figure 4.**
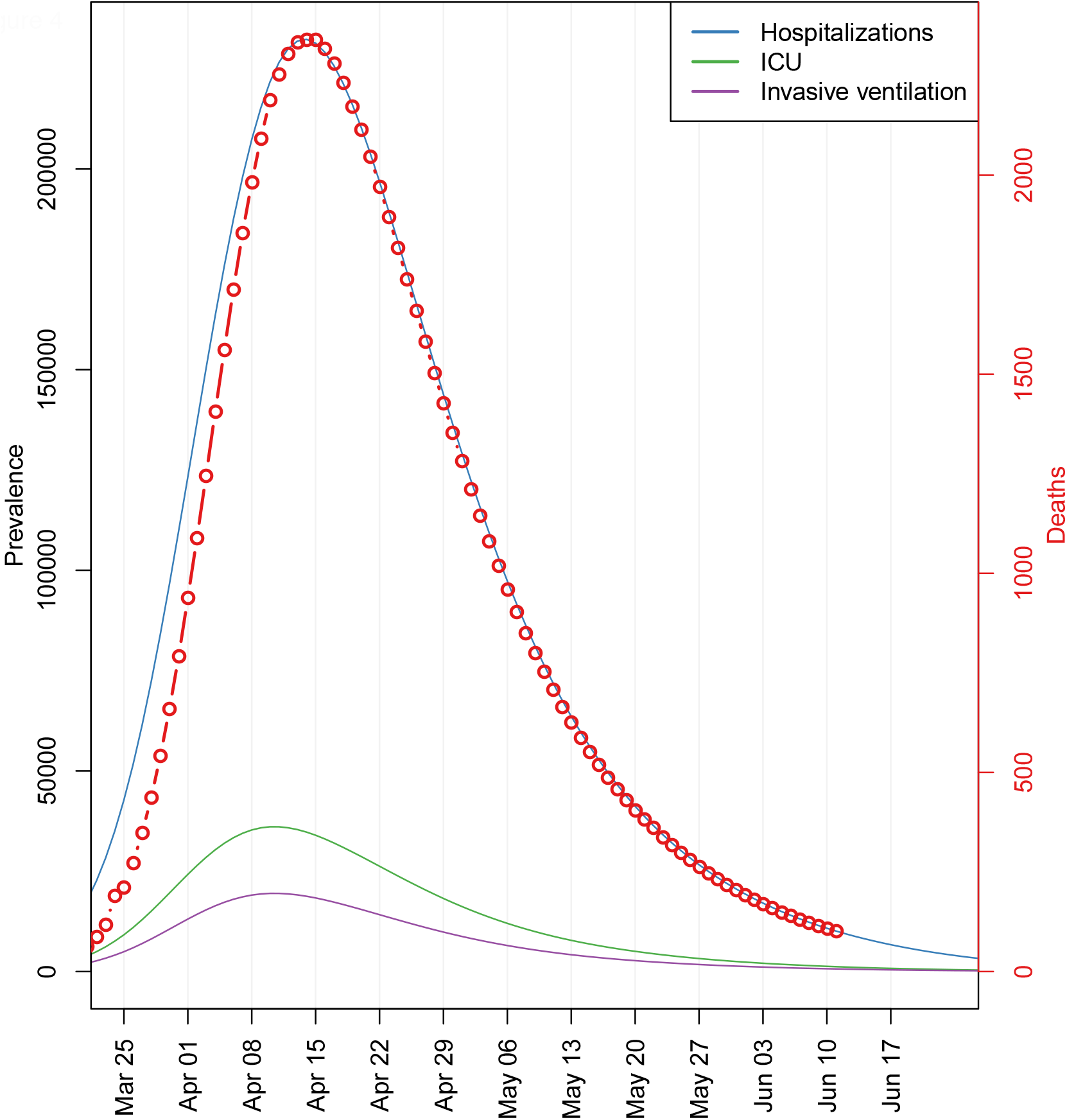
Estimates of hospitalization utilization and deaths by day, US

**Figure 5.**
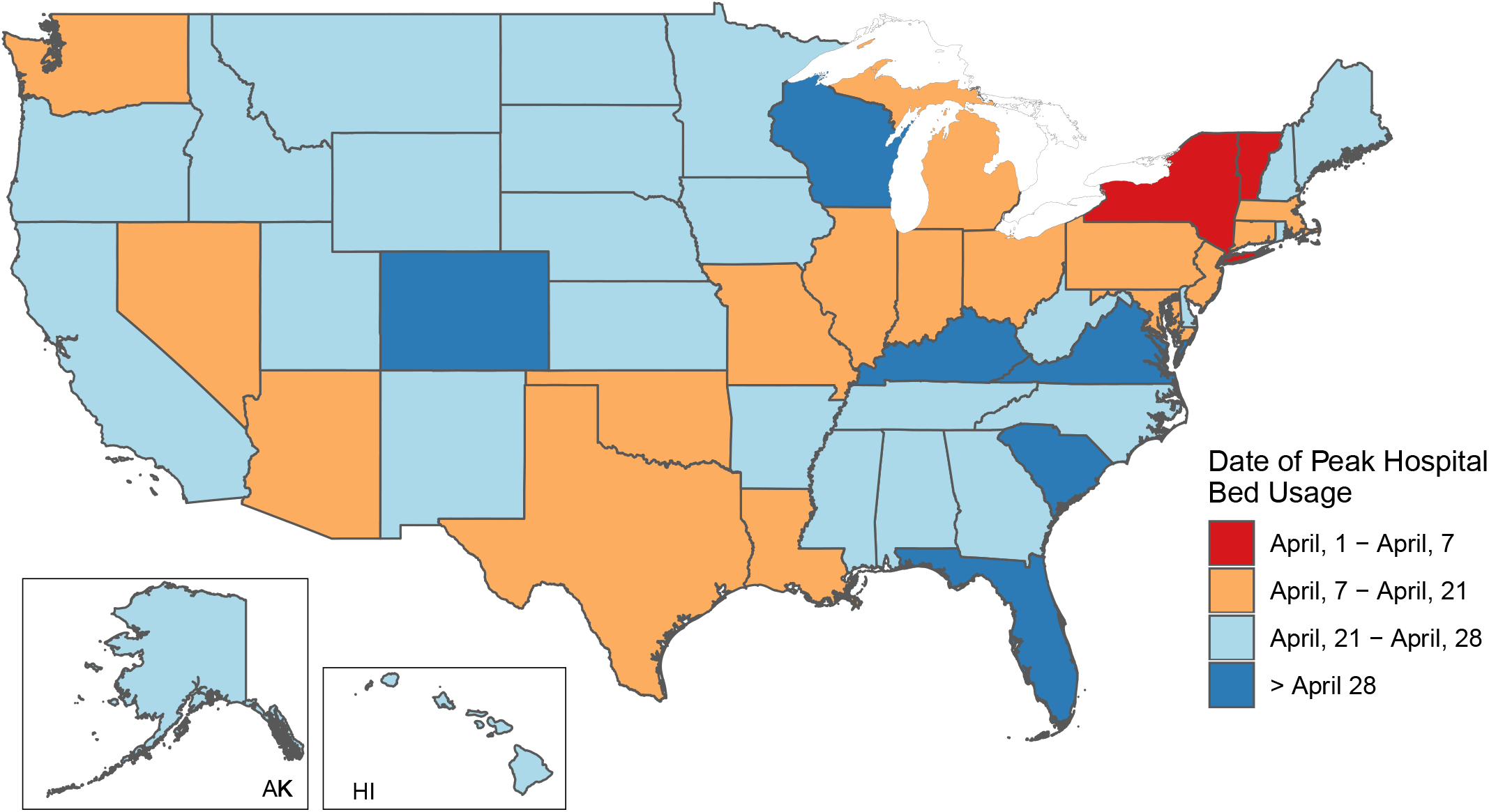
Date of peak hospital bed usage by state

Figure 6 shows aggregated excess demand for services above the capacity available currently in each state. Excess demand will be above 60,000 beds (64,175 [95% UI 7,977 to 251,059]) at the peak in the second week of April – about 7% of all hospital beds nationally. More concerning is the peak excess demand of more than 17,000 ICU beds (17,380 [95% UI 2,432 to 57,955]) – about a quarter of all ICU beds nationally. The upper bound of the 95% uncertainty intervals suggest the potential for a much more massive overload of the system, particularly amongst many patients needing ICU beds not having this level of care available. We have not been able to estimate current ventilator capacity; however, the number of ventilators implied by the peak (19,481 [95% UI 9,767 to 39,674]; Figure 4) also suggests potentially large gaps in availability of ventilators. Peak excess demand varies considerably by state; Figure 7 and 8 shows the peak % excess demand by state for total beds and ICU beds, respectively. Peak excess demand for total beds is particularly high in states such as New York, New Jersey, Connecticut, and Michigan. Peak excess demand for ICU beds is more of an issue across all states and is highest in the same set of states listed above as well as Louisiana, Missouri, Nevada, Vermont and Massachusetts.

**Figure 6.**
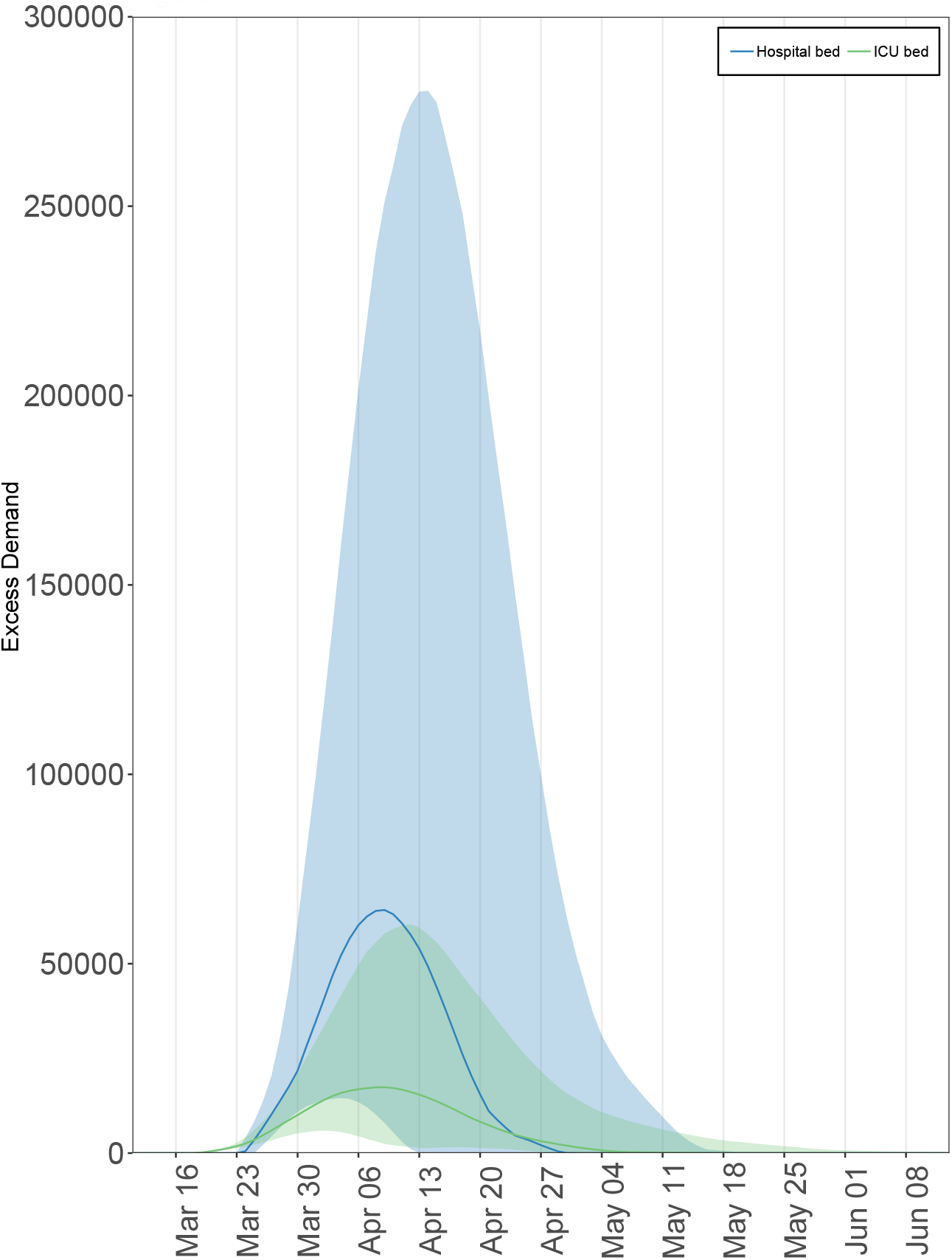
Excess demand for services above capacity available currently.

**Figure 7.**
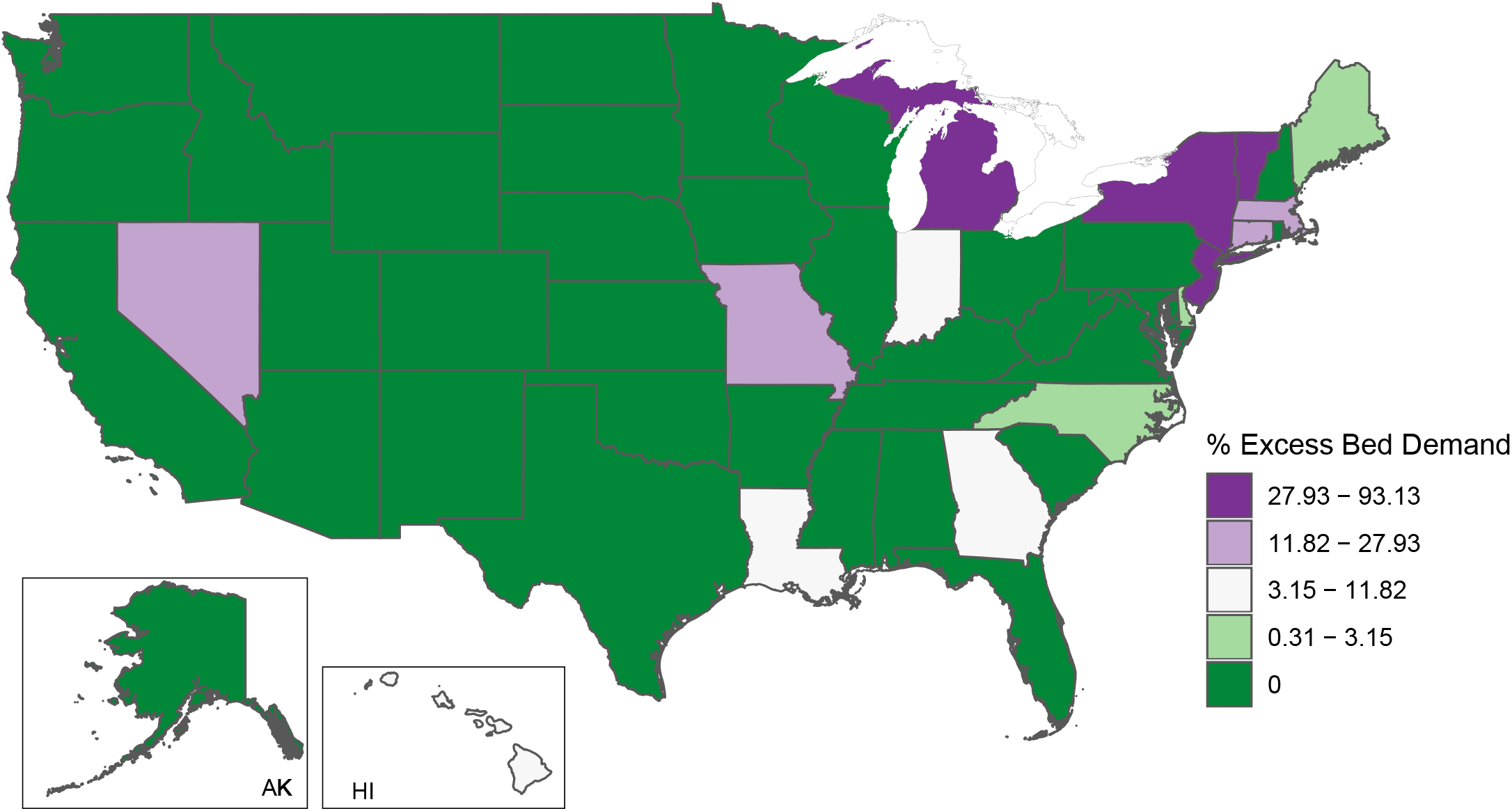
Peak % excess demand by state for total beds.

**Figure 8.**
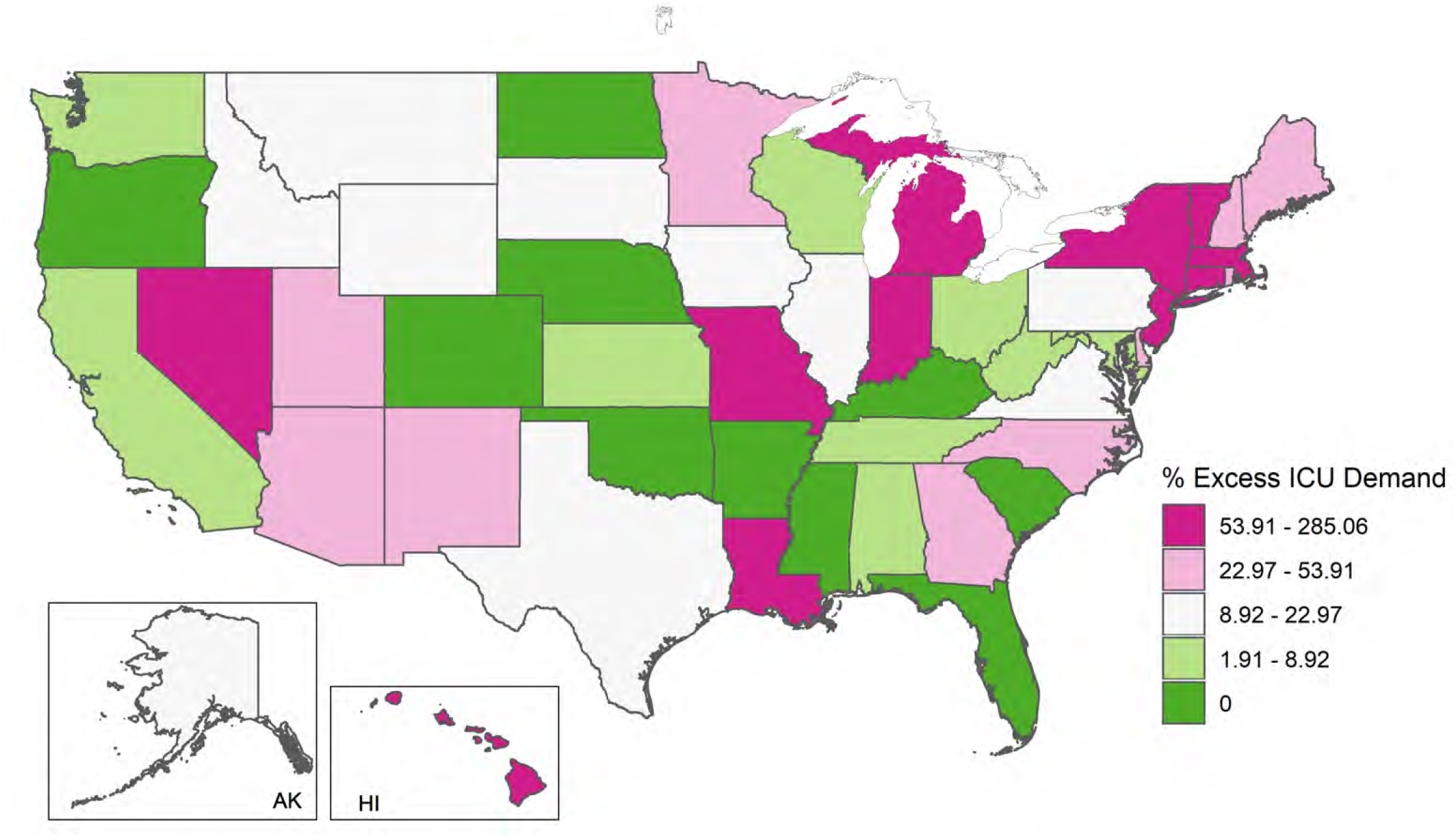
Peak % excess demand by state for ICU beds

Figure 9 shows the expected cumulative death numbers with 95% uncertainty intervals. The average forecast suggests 81,114 deaths, but the range is large, from 38,242 to 162,106 deaths. The figure shows that uncertainty widens markedly as the peak of the epidemic approaches, given that the exact timing of the peak is uncertain. Based on our projections, the number of daily deaths in the US will likely drop below 10 deaths between May 31 and June 6 (Figure 10). The date at which the projected daily death rate drops below 0.3 per million by state varies from the first half of May to the first of July (Figure 11). Those states where the death rate drops early overlap considerably with those states with large peak excess demand.

**Figure 9.**
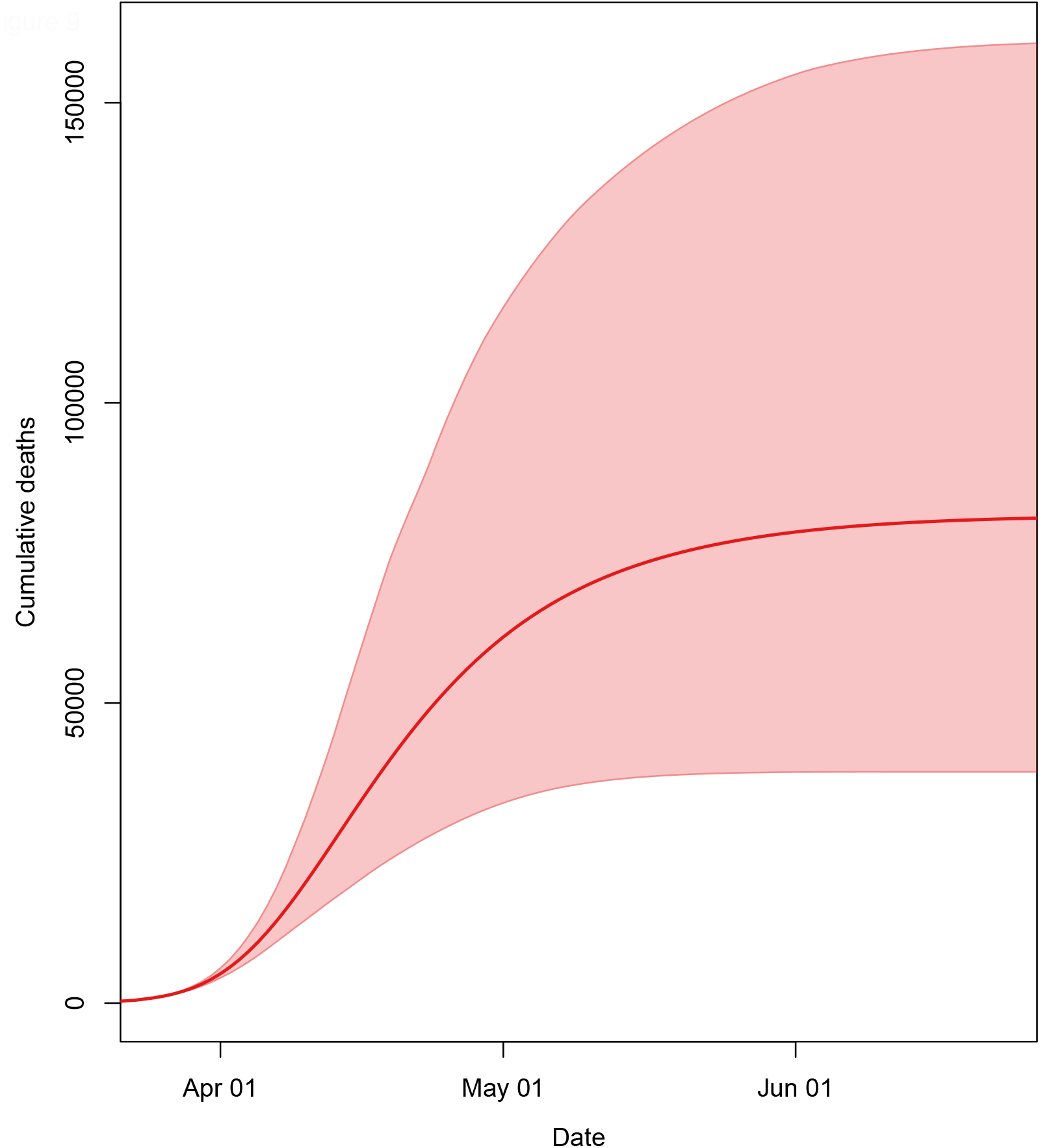
Expected cumulative death numbers with 95% uncertainty intervals

**Figure 10.**
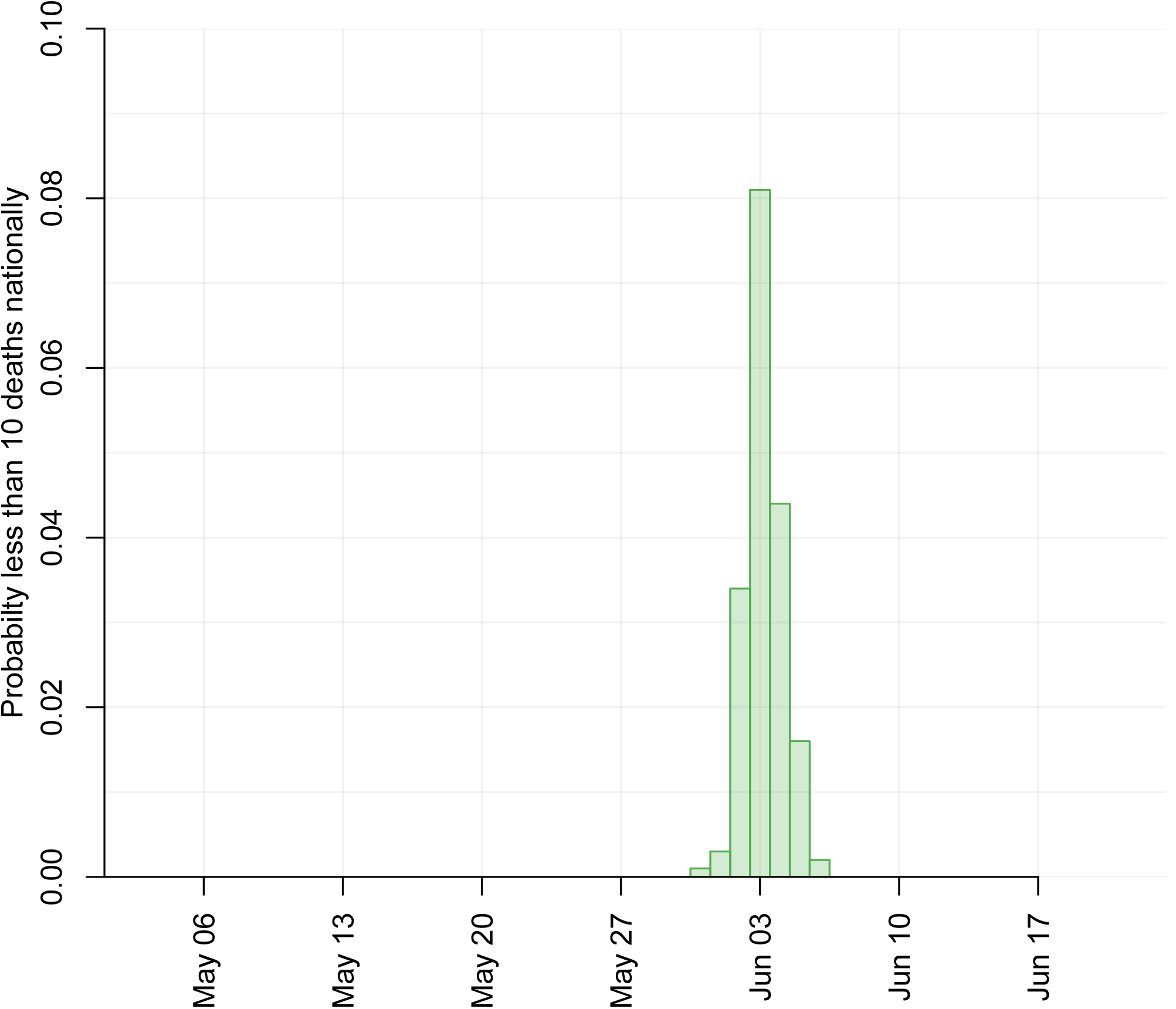
Probability by date that the number of daily deaths in the US will be below 10 deaths.

**Figure 11.**
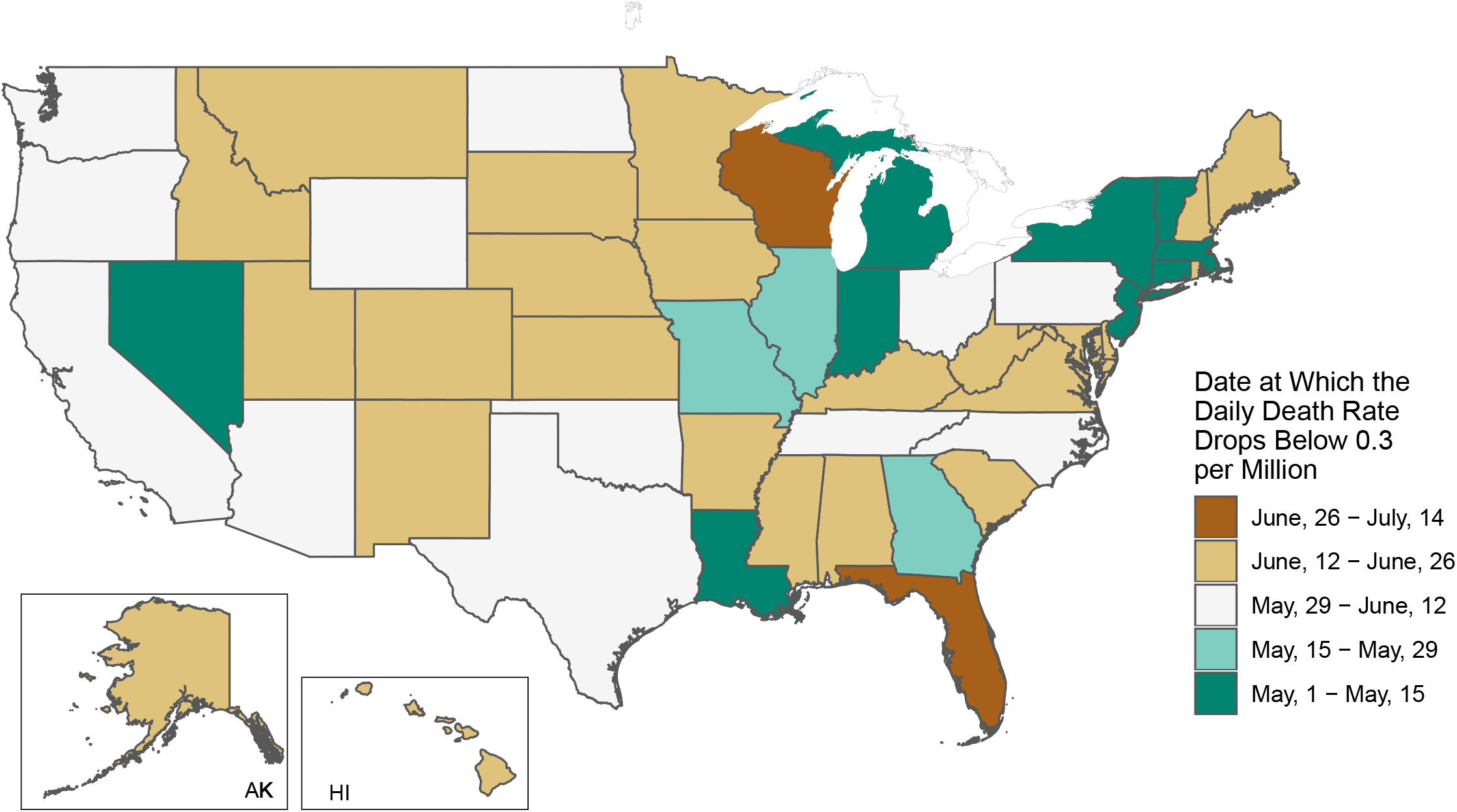
Date at which the daily death rate is projected to drop below 0.3 per million by state.

Results for each state are accessible through a visualization tool at http://covid19.healthdata.org/projections - the estimates presented in this tool will be continually updated as new data are incorporated and ultimately will supersede the results in this paper. Summary information on cumulative deaths, the date of peak demand, the peak demand, peak excess demand, and aggregate demand are provided for each US state in Table 1.

**Table 1.** Summary information on deaths, peak demand, peak excess demand, and aggregate demand, by state

| State | Cumulative Deaths | Date of Peak Hospital Use | Beds Used at Peak | ICU Beds Used at Peak | Ventilators Used at Peak | Excess Bed Demand | Excess ICU Demand | Cumulative Bed Days | Cumulative ICU Days | Cumulative Ventilator Days |
| --- | --- | --- | --- | --- | --- | --- | --- | --- | --- | --- |
| Alabama | 1155 (622–1737) | 04/24/2020 (04/24/2020–04/24/2020) | 3628 (3079–4070) | 567 (480–641) | 307 (261–347) | 0 (0–0) | 93 (6–167) | 122930 (69537–180580) | 18274 (10010–27280) | 9869 (5404–14740) |
| Alaska | 154 (62–264) | 04/26/2020 (04/27/2020–04/24/2020) | 524 (410–648) | 81 (62–102) | 44 (33–55) | 0 (0–0) | 27 (8–48) | 18417 (8104–30667) | 2725 (1138–4652) | 1473 (616–2528) |
| Arizona | 1687 (1283–2133) | 04/19/2020 (04/18/2020–04/19/2020) | 5342 (4775–5842) | 842 (752–925) | 455 (405–499) | 0 (0–0) | 334 (244–417) | 176047 (134332–221524) | 26189 (19779–33218) | 14145 (10643–17991) |
| Arkansas | 707 (367–1082) | 04/25/2020 (04/25/2020–04/27/2020) | 2183 (1841–2442) | 341 (285–385) | 184 (154–208) | 0 (0–0) | 0 (0–0) | 73840 (40262–110228) | 10982 (5809–16672) | 5930 (3137–9010) |
| California | 6109 (778–17163) | 04/24/2020 (04/09/2020–04/30/2020) | 15242 (3131–38098) | 2318 (491–5787) | 1252 (264–3114) | 0 (0–11444) | 325 (0–3794) | 651881 (84753–1807008) | 96897 (12272–271432) | 52336 (6603–146542) |
| Colorado | 940 (149–2412) | 04/28/2020 (04/16/2020–05/01/2020) | 2260 (532–4905) | 342 (81–747) | 185 (43–403) | 0 (0–54) | 0 (0–193) | 103530 (16353–262516) | 15368 (2363–39285) | 8297 (1268–21206) |
| Connecticut | 773 (250–1824) | 04/10/2020 (04/08/2020–04/14/2020) | 3341 (1266–7631) | 541 (213–1232) | 292 (114–667) | 1603 (0–5893) | 442 (114–1133) | 80434 (26752–188243) | 11971 (3886–28287) | 6465 (2096–15317) |
| Delaware | 228 (114–347) | 04/24/2020 (04/25/2020–04/24/2020) | 725 (605–809) | 113 (92–128) | 61 (50–70) | 29 (0–113) | 72 (51–87) | 23911 (12621–36026) | 3554 (1798–5475) | 1919 (965–2968) |
| District of Columbia | 132 (66–207) | 04/24/2020 (04/24/2020–04/24/2020) | 408 (339–479) | 63 (52–77) | 34 (28–42) | 0 (0–0) | 22 (11–36) | 13787 (7199–20964) | 2051 (1029–3217) | 1108 (555–1745) |
| Florida | 3342 (317–9017) | 05/14/2020 (04/20/2020–05/20/2020) | 5576 (939–14429) | 841 (142–2173) | 454 (78–1178) | 0 (0–0) | 0 (0–478) | 335283 (32340–900699) | 49981 (4716–134771) | 27002 (2532–72958) |
| Georgia | 3165 (1036–5600) | 04/21/2020 (04/13/2020–04/25/2020) | 10436 (4079–17869) | 1606 (628–2747) | 868 (338–1474) | 2114 (0–9547) | 1017 (39–2158) | 354633 (116532–624314) | 52603 (17041–93355) | 28409 (9168–50506) |
| Hawaii | 352 (168–551) | 04/25/2020 (04/25/2020–04/24/2020) | 1070 (878–1210) | 167 (134–191) | 90 (72–103) | 114 (0–254) | 122 (89–146) | 35824 (18058–54681) | 5334 (2570–8296) | 2879 (1386–4494) |
| Idaho | 388 (181–610) | 04/25/2020 (04/25/2020–04/22/2020) | 1206 (977–1374) | 188 (150–216) | 101 (80–118) | 0 (0–0) | 37 (0–65) | 41329 (20458–63118) | 6143 (2916–9566) | 3318 (1568–5173) |
| Illinois | 2454 (507–5857) | 04/16/2020 (04/09/2020–04/18/2020) | 8855 (2722–17145) | 1391 (429–2704) | 751 (230–1469) | 0 (0–2593) | 260 (0–1573) | 259637 (57195–606941) | 38607 (8170–91565) | 20845 (4382–49321) |
| Indiana | 2440 (598–7229) | 04/14/2020 (04/09/2020–04/15/2020) | 10458 (3666–30652) | 1680 (595–4993) | 907 (322–2689) | 1973 (0–22167) | 974 (0–4287) | 259565 (67206–758821) | 38578 (9630–113862) | 20833 (5199–61486) |
| Iowa | 742 (385–1125) | 04/24/2020 (04/24/2020–04/24/2020) | 2250 (1922–2517) | 353 (298–397) | 190 (161–215) | 0 (0–0) | 107 (52–151) | 75241 (40903–111618) | 11216 (5907–16946) | 6058 (3187–9165) |
| Kansas | 669 (329–1044) | 04/24/2020 (04/25/2020–04/25/2020) | 2036 (1680–2293) | 317 (256–362) | 172 (138–196) | 0 (0–0) | 39 (0–84) | 69582 (35854–106570) | 10354 (5112–16141) | 5592 (2757–8722) |
| Kentucky | 585 (63–1646) | 05/05/2020 (04/14/2020–05/10/2020) | 1172 (215–3023) | 176 (32–465) | 95 (18–251) | 0 (0–0) | 0 (0–17) | 62720 (6799–174924) | 9320 (978–26205) | 5033 (522–14158) |
| Louisiana | 2081 (946–5486) | 04/08/2020 (04/04/2020–04/14/2020) | 9217 (4778–23322) | 1484 (787–3715) | 801 (425–2010) | 2013 (0–16118) | 1007 (310–3238) | 223578 (101330–586716) | 33228 (15070–87685) | 17942 (8143–47252) |
| Maine | 334 (174–511) | 04/24/2020 (04/24/2020–04/23/2020) | 1086 (918–1218) | 170 (140–194) | 92 (75–105) | 25 (0–157) | 106 (76–130) | 34909 (19103–51943) | 5194 (2734–7873) | 2804 (1473–4259) |
| Maryland | 857 (74–3381) | 04/19/2020 (04/16/2020–04/14/2020) | 2232 (457–7129) | 345 (66–1150) | 186 (35–624) | 0 (0–3168) | 79 (0–884) | 92613 (8920–356207) | 13755 (1209–53899) | 7428 (653–29144) |
| Massachusetts | 2231 (363–7089) | 04/12/2020 (04/09/2020–04/15/2020) | 9857 (2084–28403) | 1606 (353–4606) | 867 (188–2486) | 5009 (0–23555) | 1329 (76–4329) | 232779 (40191–723480) | 34637 (5739–109258) | 18705 (3091–59024) |
| Michigan | 4061 (1361–10955) | 04/08/2020 (04/06/2020–04/09/2020) | 20717 (9152–52023) | 3473 (1549–8884) | 1876 (842–4767) | 10563 (0–41869) | 2731 (807–8142) | 425999 (152563–1120612) | 63352 (21765–169288) | 34204 (11753–91393) |
| Minnesota | 1280 (697–1916) | 04/24/2020 (04/23/2020–04/25/2020) | 3973 (3393–4463) | 621 (524–702) | 336 (283–379) | 0 (0–0) | 266 (169–347) | 134756 (76858–197954) | 20043 (11113–29825) | 10827 (5988–16133) |
| Mississippi | 675 (380–1003) | 04/23/2020 (04/23/2020–04/23/2020) | 2141 (1842–2407) | 334 (284–381) | 181 (152–206) | 0 (0–0) | 0 (0–41) | 72166 (41957–105077) | 10722 (6057–15856) | 5790 (3261–8581) |
| Missouri | 2977 (384–9636) | 04/20/2020 (04/13/2020–04/22/2020) | 10630 (2339–31875) | 1669 (366–5041) | 901 (197–2733) | 2697 (0–23942) | 1111 (0–4483) | 311155 (43337–992220) | 46290 (6131–148916) | 24999 (3307–80425) |
| Montana | 251 (108–402) | 04/25/2020 (04/26/2020–04/27/2020) | 784 (598–898) | 122 (91–143) | 66 (49–78) | 0 (0–0) | 37 (6–58) | 26198 (12146–41425) | 3900 (1729–6295) | 2106 (934–3409) |
| Nebraska | 437 (200–699) | 04/25/2020 (04/25/2020–04/26/2020) | 1323 (1068–1504) | 206 (162–238) | 111 (87–129) | 0 (0–0) | 0 (0–6) | 45184 (22080–70619) | 6724 (3165–10684) | 3631 (1708–5786) |
| Nevada | 801 (120–2585) | 04/15/2020 (04/10/2020–04/18/2020) | 3515 (715–10939) | 563 (120–1759) | 304 (64–953) | 1268 (0–8692) | 380 (0–1576) | 89067 (14114–281580) | 13214 (2022–42379) | 7136 (1087–22925) |
| New Hampshire | 318 (155–496) | 04/24/2020 (04/25/2020–04/25/2020) | 1027 (849–1164) | 161 (128–184) | 87 (69–100) | 9 (0–146) | 78 (45–101) | 34260 (17549–51898) | 5090 (2498–7857) | 2750 (1345–4257) |
| New Jersey | 4109 (1608–10409) | 04/11/2020 (04/06/2020–04/14/2020) | 17644 (8822–42408) | 2836 (1463–6876) | 1531 (787–3692) | 9829 (1007–34593) | 2371 (998–6411) | 430475 (173471–1080824) | 64063 (25253–162147) | 34584 (13601–87453) |
| New Mexico | 513 (245–803) | 04/25/2020 (04/25/2020–04/24/2020) | 1587 (1313–1783) | 247 (200–282) | 134 (107–152) | 0 (0–31) | 130 (83–165) | 53736 (27359–81727) | 7993 (3900–12398) | 4315 (2108–6695) |
| New York | 10243 (4911–26983) | 04/06/2020 (04/03/2020–04/11/2020) | 48311 (26865–122319) | 7972 (4489–19893) | 4305 (2459–10757) | 35301 (13855–109309) | 7254 (3771–19175) | 1057788 (514026–2774647) | 157459 (75373–414490) | 85028 (40771–224139) |
| North Carolina | 2411 (1600–3291) | 04/22/2020 (04/20/2020–04/22/2020) | 7811 (6908–8644) | 1224 (1086–1360) | 661 (584–735) | 686 (0–1519) | 657 (519–793) | 259776 (176831–350701) | 38593 (25905–52522) | 20839 (13975–28410) |
| North Dakota | 163 (72–265) | 04/25/2020 (04/26/2020–04/24/2020) | 495 (388–568) | 77 (59–91) | 42 (32–50) | 0 (0–0) | 0 (0–5) | 16455 (7560–26041) | 2454 (1068–3977) | 1327 (573–2161) |
| Ohio | 2733 (2066–3460) | 04/16/2020 (04/15/2020–04/16/2020) | 8759 (7858–9657) | 1381 (1236–1530) | 746 (666–827) | 0 (0–0) | 143 (0–292) | 285835 (217010–361279) | 42521 (32020–54219) | 22962 (17254–29328) |
| Oklahoma | 898 (669–1147) | 04/17/2020 (04/16/2020–04/18/2020) | 2873 (2548–3179) | 452 (401–500) | 244 (216–271) | 0 (0–0) | 0 (0–33) | 94765 (70892–120433) | 14090 (10426–18081) | 7609 (5610–9791) |
| Oregon | 584 (91–1378) | 04/24/2020 (04/08/2020–04/28/2020) | 1462 (352–3047) | 223 (55–466) | 120 (30–252) | 0 (0–390) | 13 (0–256) | 61463 (9509–143860) | 9139 (1372–21595) | 4935 (737–11666) |
| Pennsylvania | 3094 (2356–3892) | 04/17/2020 (04/16/2020–04/17/2020) | 9745 (8700–10642) | 1535 (1370–1687) | 829 (738–911) | 0 (0–0) | 492 (327–644) | 316655 (241507–397108) | 47155 (35627–59710) | 25475 (19191–32383) |
| Rhode Island | 245 (125–373) | 04/24/2020 (04/24/2020–04/23/2020) | 769 (641–869) | 120 (98–138) | 65 (52–75) | 0 (0–74) | 79 (57–97) | 25360 (13488–37901) | 3776 (1927–5770) | 2039 (1047–3120) |
| South Carolina | 768 (85–2028) | 05/02/2020 (04/13/2020–05/06/2020) | 1697 (294–3804) | 256 (46–577) | 139 (25–312) | 0 (0–0) | 0 (0–173) | 82227 (8922–214743) | 12216 (1306–32153) | 6596 (702–17398) |
| South Dakota | 201 (83–323) | 04/26/2020 (04/26/2020–04/25/2020) | 606 (473–690) | 95 (71–110) | 51 (38–60) | 0 (0–0) | 21 (0–36) | 20364 (9108–32358) | 3031 (1281–4946) | 1636 (687–2694) |
| Tennessee | 1551 (1069–2083) | 04/21/2020 (04/21/2020–04/22/2020) | 4995 (4399–5511) | 783 (692–865) | 423 (373–469) | 0 (0–0) | 154 (63–236) | 165849 (116021–219815) | 24645 (16994–32967) | 13308 (9170–17834) |
| Texas | 5847 (4249–7668) | 04/18/2020 (04/18/2020–04/18/2020) | 19135 (16951–21851) | 2986 (2627–3465) | 1612 (1408–1864) | 0 (0–0) | 727 (368–1206) | 648770 (472173–848002) | 96270 (69161–127252) | 51986 (37159–68919) |
| Utah | 619 (335–958) | 04/24/2020 (04/24/2020–04/22/2020) | 1958 (1656–2374) | 305 (253–375) | 165 (137–203) | 0 (0–0) | 135 (83–205) | 68354 (38440–104047) | 10143 (5537–15686) | 5478 (2981–8498) |
| Vermont | 386 (146–822) | 04/05/2020 (03/31/2020–04/07/2020) | 1953 (966–3876) | 325 (173–657) | 175 (94–355) | 1420 (433–3343) | 291 (139–623) | 40503 (15971–83249) | 6023 (2320–12667) | 3252 (1252–6807) |
| Virginia | 1543 (152–4131) | 05/02/2020 (04/13/2020–05/05/2020) | 3435 (557–7932) | 519 (85–1210) | 280 (46–654) | 0 (0–1351) | 190 (0–881) | 167373 (16748–443608) | 24860 (2426–66459) | 13424 (1306–35934) |
| Washington | 1429 (312–2710) | 04/19/2020 (03/27/2020–04/23/2020) | 2922 (944–5546) | 440 (147–841) | 238 (79–456) | 0 (0–639) | 99 (0–500) | 154707 (32073–293961) | 22987 (4705–44001) | 12415 (2531–23819) |
| West Virginia | 460 (193–774) | 04/26/2020 (04/26/2020–04/23/2020) | 1402 (1078–1618) | 218 (161–255) | 118 (87–138) | 0 (0–0) | 22 (0–59) | 47825 (21187–77858) | 7117 (2983–11787) | 3844 (1605–6363) |
| Wisconsin | 853 (106–1993) | 05/22/2020 (04/23/2020–05/23/2020) | 1358 (301–3137) | 204 (45–471) | 110 (24–256) | 0 (0–0) | 32 (0–299) | 89330 (10867–208544) | 13284 (1567–31212) | 7174 (833–16899) |
| Wyoming | 136 (59–224) | 04/26/2020 (04/26/2020–04/25/2020) | 436 (339–505) | 68 (52–82) | 37 (28–44) | 0 (0–0) | 24 (8–38) | 14604 (6804–23575) | 2167 (963–3598) | 1170 (513–1952) |

## Discussion

This study has generated the first set of estimates of predicted health service utilization and deaths due to COVID-19 by day for the next 4 months for all US states, assuming that social distancing efforts will continue throughout the epidemic. The analysis shows large gaps between need for hospital services and available capacity, especially for inpatient and ICU beds. A similar or perhaps even greater gap for ventilators is also likely, but detailed state data on ventilator capacity is not available to directly estimate that gap. Uncertainty in the time course of the epidemic, its duration, and the peak of utilization and deaths is large this early in the epidemic. Given this, it is critical to update these projections as new data on deaths in the US are collected. Uncertainty will also be reduced as we gain more knowledge about the course of the epidemic in other countries, particularly in Europe, where countries such as Italy and Spain have a more advanced epidemic than the US. A critical aspect to the size of the peak is when aggressive measures for social distancing are implemented in each state. Delays in implementing government-mandated social distancing will have an important effect on the resource gaps that states’ health systems will have to manage.

Our estimates of excess demand suggest hospital systems will face difficult choices to continue providing high-quality care to their patients in need. This model was first developed for use by the UW Medicine system, and their practical experience provides insight into how it may be useful for planning purposes. From the perspective of planning for the 4 hospitals in the UW Medicine system, these projections immediately made apparent the need to rapidly build internal capacity. Strategies to do so included the suspension of elective and non-urgent surgeries and procedures, while supporting surge planning efforts and reconfiguration of medical/surgical and ICU beds across the system. This information also allowed us to more effectively engage UW Medicine leadership in clinician and staff workforce pooling and redeployment planning and trigger points while focusing efforts to seek and secure necessary personal protective equipment (PPE) and other equipment to close the identified gap. It also supported a proactive discussion regarding the potential shift from current standards of care to crisis standards of care, with the goal to do the most good for the greatest number in the setting of limited resources.

There are a variety of options available to deal with the situation, some of which have already been implemented or are being implemented in Washington and New York. One option is to reduce non-COVID-19 patient use. State governments have cancelled elective procedures^38–40^ (and many hospitals but not all have followed suit). However, this decision has significant financial implications for health systems, as elective procedures are a major source of revenue for hospitals.^41^ For example, in the UW Medicine system, non-COVID-19 utilization is down 14% over a 2-week period since elective activity was reduced. Also, aggressive social distancing policies reduce not only the transmission of COVID-19 but will likely have the added benefit of reducing health care utilization due to other causes such as injuries. Reducing non-COVID-19 demand alone will not be sufficient, and strategies to increase capacity are clearly needed. This includes setting up additional beds by repurposing unused operating rooms, pre- and post-recovery rooms, procedural areas, medical and nursing staff quarters, and hallways. For example, in UW Medicine, the use of such strategies has enabled planning to increase bed capacity temporarily by 65%.

Currently, one of the largest constraints on effective care may be the lack of ventilators. One supplement to ventilator capacity is using anesthesia machines freed up by deferring or cancelling elective surgeries. Other options go beyond the capacity or control of specific hospitals. The use of mobile military resources including the National Guard^42–44^ has the potential to address some capacity limitations, particularly given the differently timed epidemics across states. Other innovative strategies will need to be found, including the construction of temporary hospital facilities as was done in Wuhan,^45^ Washington state,^46^ and also New York.^44,47^

In this study, we have quantified the potential gap in physical resources, but there is an even larger potential gap in human resources (HR). Expanding bed capacity beyond licensed bed capacity may require an even larger increase in the HR to provide care. The average annual bed- day utilization rate in the US is 66% and ranges from 54% (Idaho) to 80% (Connecticut) by state. Most US hospitals are staffed appropriately at their usual capacity utilization rate, and expanding even up to, but then potentially well beyond, licensed capacity will require finding substantial additional HR. Strategies include increasing overtime, training operating room and community clinic staff in inpatient care or physician specialties in COVID-19 patient care, rehiring recently separated workers, and the use of volunteers. For example, UW Medicine has been fortunate that clinical faculty time can be redirected from research and teaching to clinical care during the COVID-19 surge. Other hospitals may not have this same ability. The most concerning HR bottleneck identified for UW Medicine is for ICU nurses, for which there are very limited options for increasing capacity. In addition to HR, what should not be overlooked is the increased demand for supplies ranging from PPE, medication, and ventilator supplies to basics such as bed linen. Add to these the need to expand other infrastructure required to meet the COVID-19 surge, such as information technology (IT) for electronic medical records. The overall financial cost over a short period of time is likely to be enormous, particularly when juxtaposed against the substantial reductions in revenue due to the cancellation of elective procedures.

The timing of the implementation of social distancing mandates may be a critical determinant of peak demand and cumulative deaths. For states that have not implemented 3 of 4 measures (school closures, closing non-essential services, shelter-in-place, and major travel restrictions), we have assumed that they will be implemented within 7 days, given the rapid adoption of these measures in nearly all states. At this point in the epidemic, we have had to make arbitrary assumptions in our model on the equivalency between implementing 1, 2, or 3 measures – and we have implicitly assumed that implementing 3 of 4 measures will be enough to follow a trajectory similar to Wuhan – but it is plausible that it requires all 4 measures. As more data accumulate, especially on the timing of deceleration of daily deaths, we may be able to empirically test which of these measures is more correlated with slowing the epidemic curve and reducing the ultimate death toll. Perhaps as important will be the question of adherence to social distancing mandates; it will take time to evaluate whether social distancing adherence is fundamentally different in the US compared to Wuhan. Even in Wuhan it was a full 27 days from implementation of social distancing to reaching the peak level of daily deaths.

As data on the epidemics in each admin 1 unit accumulate, including data on health service utilization, we will derive important insights into the epidemic trajectories and health service demand. At this early stage, even 1 to 2 days’ more data for a state will improve the estimates of service need and expected deaths. Because of sparse US data for some aspects of health service utilization, we have used data from the US, Italy, and China. As more data on US treatment accumulate, further revisions will be able to more accurately reflect US practice patterns for COVID-19. For this reason, we will revise the model every day, providing an updated forecast for health service providers and the public.

Any attempt to forecast the COVID-19 epidemic has many limitations. Only one location has had a generalized epidemic and has currently brought new cases to 0 or near 0, namely Wuhan. Many other locations, including all other provinces in China, have so far successfully contained transmission, preventing a general outbreak. Modeling for US states based on one completed epidemic, at least for the first wave, and many incomplete epidemics is intrinsically challenging. The consequent main limitation of our study is that observed epidemic curves for COVID-19 deaths define the likely trajectory for US states. In this study, we do include a covariate meant to capture the timing of social distancing measures to take into account that Wuhan implemented 4 out of 4 social distancing measures within 6 days of reaching a threshold death rate of 0.31 per million. Our models explicitly take into account variation in age-structure, which is a key driver of all-age mortality. But these efforts at quantification do not take into account many other factors that may influence the epidemic trajectory: the prevalence of chronic lung disease, the prevalence of multi-morbidity, population density, use of public transport, and other factors that may influence the immune response. We also have not explicitly incorporated the effect of reduced quality of care due to stressed and overloaded health systems beyond what is captured in the data. For example, the higher mortality rate in Italy is likely in part due to policies around restricting invasive ventilation in the elderly. The model ensemble used does suggest that locations with faster increases in the death rate are likely to have more peak case load and cumulative deaths, but our uncertainty intervals are appropriately large.

## Conclusion

COVID-19 is an extraordinary challenge to US health and the healthcare system. In this study, we forecast a huge excess of demand for hospital bed-days and ICU bed-days, especially in the second week of April. Our estimate of 81 thousand deaths in the US over the next 4 months is an alarming number, but this number could be substantially higher if excess demand for health system resources is not addressed and if social distancing policies are not vigorously implemented and enforced across all states. This planning model will hopefully provide an up-to-date tool for improved hospital resource allocation.

## Data Availability

A full list of data citations are available by contacting the corresponding author.

